# Predicting Transdiagnostic Social Impairments in Childhood using Connectome-based Predictive Modeling

**DOI:** 10.1101/2022.04.07.22273518

**Authors:** Alexander J. Dufford, Violet Kimble, Link Tejavibulya, Javid Dadashkarimi, Karim Ibrahim, Denis G. Sukhodolsky, Dustin Scheinost

## Abstract

**Background:** Social impairments are core features of multiple neurodevelopmental disorders. Previous neuroimaging studies have focused on elucidating associations between brain function and social impairments within disorders but have not predicted these impairments from brain connectivity in a transdiagnostic manner, across several diagnostic categories. This study used a machine learning approach to examine functional connectivity that predicts elevated social impairments in a transdiagnostic sample of youths. We hypothesized that predictive edges would be from brain regions involved in social cognition.

**Methods:** Connectome-based predictive modeling (CPM) was used to build a transdiagnostic model of social impairments as measured by the Social Responsiveness Scale (SRS-2, raw score >75). We used functional connectivity data during a social movie-watching task from the Healthy Brain Network data (N=144, mean age=11.68 (3.52), 32% male). The average number of diagnoses was 3.4 (SD = 1.82, range = 0–11), including ASD (40.9%), ADHD (79%), mood disorders (15.9%), and anxiety disorders (43%). A similar transdiagnostic sample high SRS-2 scores (n=41) was used for replication.

**Results:** SRS-2 scores were predicted from functional connectivity data using both 10-fold cross-validation (median *q*^*2*^=0.32, *r*=0.57, *p*<.001) and leave-one-group-out cross-validation (median *q*^*2*^*’s*>0.04, *r’s*>0.36, *p’s*<.001). Predictive connections were widely distributed across the brain but were rooted in regions involved in social cognition, the subcortex, and the salience network. The model successfully predicted SRS-2 scores in the replication sample (*r=*0.33, *p*<.035, *df*=39).

**Conclusion:** We identified connectivity patterns predictive of social impairments in a transdiagnostic sample. These networks have the potential to provide insight into development novel targeted interventions for social impairments across traditional diagnostic categories.

## Introduction

Social impairments are common features of multiple neurodevelopmental disorders including autism spectrum disorder (ASD) and attention-deficit/hyperactivity disorder (ADHD) [1, 2]. To better understand these common features, the Research Domain Criteria (RDoC) has provided a fully dimensional alternative to more traditional categorically-based diagnostic systems [3, 4]. For example, ASD and ADHD are defined as distinct diagnoses with substantially overlapping clinical presentations [5, 6], including social impairment. Focusing on social impairments by only one disorder at a time limits the comprehensiveness of these brain-phenotype associations and by fails to capture the commonalities and dimensional aspects of the behavioral presentations and underlying neurobiology.

The potential utility of neuroimaging for identifying transdiagnostic brain markers of social impairments is strong [7-13]. Yet, recent literature has found it challenging to find converging evidence from this body of research [14-16], noting issues with replication [17-20]. Potentially, two factors contribute to this. First, studies have focused on one disorder, lacking transdiagnostic (i.e., dimensional) approaches to social impairments. Transdiagnostic models putatively find more generalizable brain features underlying behavior—rather than idiosyncratic ones to a specific disorder. Second, these studies have been limited in their ability to ‘predict’ impairments in novel individuals. It remains critical to test for out-of-sample replication as only examining associations within a single sample will inflate effects sizes.

Connectome-based predictive modeling (CPM) is a well-validated machine learning approach to building predictive models of transdiagnostic social impairments from whole-brain functional connectivity data, or ‘connectomes’ [21]. Compared to explanatory models using correlation or regression, CPM protects against overfitting by being trained and validated in independent samples. CPM has been shown to be a useful analytic, data-driven framework for both transdiagnostic prediction and pediatric studies [7]. For example, it was shown that CPM models of Social Responsiveness Scale (SRS-2) built o-n children with ASD also predicted ADHD Rating Scale-IV in children with ADHD. Thus, CPM has the potential to identify brain ‘signatures’ of brain-phenotype associations, such as social impairments, which generalize across different disorders and measures [21]. However, it remains unclear if CPM can be used to predict social impairments in a transdiagnostic sample.

In the current study, we used whole-brain functional connectomes during a movie-watching task to predict social impairments in a transdiagnostic sample of children (n=144) from the Healthy Brain Network (HBN) dataset. CPM was used to identify a set of edges (functional connections between pairs of nodes) that predicted social impairments as measured by the Social Responsiveness Scale (SRS-2). We hypothesized that there would be a high degree of overlap between these edges and the ‘social brain’. These areas include the medial prefrontal cortex, temporoparietal junction, anterior temporal lobes, inferior frontal gyrus, and posterior superior temporal sulcus [22, 23]. Next, we show that overall connectivity in these identified edges showed significant differences between children exhibiting social impairments (n=144) compared to those without impairments (n=699). Finally, to assess the generalizability of the connectome-based predictive model of SRS-2 scores, we applied the model trained on the HBN dataset (our “discovery” sample) in a separate independent transdiagnostic sample (n=41).

## Materials and Methods

### Discovery Sample

We used a large, transdiagnostic neuroimaging sample, the Child Mind Institute’s Healthy Brain Network data [24], as the “discovery” sample. As the current study was interested in social impairments, we only included individuals with high SRS-2 raw scores above a cutoff of >75 for their SRS-2 total raw score [5, 25]. Using a cutoff of a raw score of 75 has been shown to have high sensitivity and specificity (0.85 and 0.75 respectively) [25]. After excluding individuals for missing data, excessive motion during fMRI (> 0.25 mm), usable structural MRI, and an SRS-2 total score above 75, 144 participants were included for analyses. Of these participants, 59 had a diagnosis of ASD, 114 had a diagnosis of ADHD, 23 had a diagnosis of a mood disorder (Major Depressive Disorder, Persistent Depressive Disorder, Disruptive Mood Dysregulation Disorder, 63 had an anxiety disorder diagnosis, 104 had a diagnosis of ‘Other’ that did not fall into the previous categories (ASD, ADHD, mood, or anxiety), and 4 participants had no diagnosis. Co-morbidity in the sample was high with the average number of diagnoses at 3.4 (SD = 1.82, range = 0–11). Therefore, the sample was well-suited to examine transdiagnostic functional connectivity.

In addition, 699 participants from the HBN that met quality control (no missing data, motion during fMRI <0.25 mm, and usable structural MRI) and had an SRS-2 score < 75 were used to compare network strength in the network identified by CPM.

### Replication Sample

The replication sample consisted of 41 children with high SRS-2 scores (raw score > 75 to remain consistent with the discovery sample) from a previous transdiagnostic CPM study of aggression (see Supplement) [26]. Connectivity data was processed in the same manner as the discovery sample. Full detail of this sample can be found in the Supplement. The diagnostic characteristics are presented in **Table S1**.

### Social Responsiveness Scale-2 (SRS-2)

The SRS-2 has been shown to measure impairments in infants and children effectively and captures the social constructs: attachment and affiliation, social communication, and understanding of mental states [27].

### HBN Data Acquisition and Connectivity Processing

The imaging procedures and sequences for the HBN datasets have been previously reported [24]. We calculated whole-brain functional connectome for the discovery dataset (HBN) during a movie-watching task. We selected this task due to the social nature of the movie (Despicable Me). Previous work suggests that naturalistic imaging paradigms (such as movies) are better at elucidating individual differences in brain connectivity [28]. As described previously, we followed a standard preprocessing pipeline for the whole-brain functional connectomes (see Supplement for full detail).

### Predictive Modeling Framework

The study used CPM [21], a data-driven machine learning algorithm for making predictions based upon functional connectivity data. The CPM procedures used the following steps: 1) linear regression was used as feature selection to relate each edge (functional connection) in the connectome to the phenotypic variable of interest (SRS-2 score) in the training set. 2) Feature summarization was performed by summing the strength of all selected edges in the first step, which created a single summary value for each participant. 3). The model was built using linear associations between the single-participant summary value (the independent variable) and the phenotypic variable (SRS-2). Once the model was built from the training data, it was used to predict behavioral scores from connectomes of previously unseen participants taken from a testing set. Training in one dataset and performing prediction in a novel dataset demonstrates that predictive models are not simply overfitting a specific set of data; rather that they will generalize and likely replicate in future studies. A feature selection threshold of *p*<.005 was adopted for the current study. Covariates included in the model and SRS-2 total scores were participant age, sex, head motion, site, and IQ.

### Model Validation

The current study utilized four model validation approaches: 10-fold cross-validation, leave-one-group-out cross-validation, leave-one-site-out cross-validation (see Supplement), and external validation. First, the 10-fold cross-validation approach was used to train the transdiagnostic model of social impairments (SRS-2 scores). For this procedure, the entire sample (n=144 was randomly divided into 10 approximately equal groups regardless of diagnosis. The transdiagnostic model was trained on 9 groups for each fold and tested on the “left-out” 10^th^ group. Second, to demonstrate that results were not driven by better prediction in one diagnostic group, we conducted a leave-one-group-out cross-validation, which involves training the model on all but one diagnostic group and testing in the left-out diagnostic group. This model validation procedure was not conducted with multiple iterations as the possibilities for iteration are exhausted with one set.

### Prediction Performance Assessment

The model prediction performance of the 10-fold cross-validation was evaluated using the cross-validated coefficient of determination: *q*^*2*^. We calculated *q*^*2*^ using the equation: 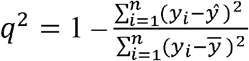. The manuscript reports the median *q*^*2*^ of the 100 iterations of 10-fold cross-validation. The significance of *q*^*2*^ was calculated via permutation testing. The correspondence between the connectivity matrices and SRS-2 scores was randomly shuffled 1000 times, and the CPM analysis was conducted again to calculate null distributions of *q*^*2*^. For convenience, Pearson’s correlation (*r*) and Spearman’s rank correlation (*ρ*) are also reported.

### Localizing the Predictive Networks

Functional networks that are predictive of a behavioral score, as calculated by CPM, are typically complex and widespread, including multiple networks and brain regions. CPM studies have previously summarized these networks at multiple levels, including the edge, node, and network levels. For the localization of predictive networks, nodes were characterized based upon their ‘degree’ or the sum of the number of edges for each node belonging to the predictive networks.

### External Replication and Validation

We followed the external replication and validation procedure previously described in other CPM studies. We examined the generalizability of the findings by testing if the CPM built in the discovery significantly predicted SRS-2 scores in the independent replication sample. As with previous cross-dataset analyses for CPM, cross-dataset model performance was assessed via the Pearson correlation between the predicted and actual scores and parametric *p*-value.

### Data Availability

This HBN data was accessed via the Child Mind Institute Biobank, Healthy Brain Network: http://fcon_1000.projects.nitrc.org/indi/cmi_healthy_brain_network/index.html in which a data usage agreement is required. This manuscript reflects the views of the authors and does not necessarily reflect the opinions or views of the Child Mind Institute.

## Results

### Sample Characteristics

Demographic information for the HBN participants is shown in **Table 1** (tables generated with the R package “scipub” https://github.com/dpagliaccio/scipub), including information regarding the SRS-2 scores for the sample. The clinical characteristics of the HBN sample are presented in **Table 2**. The demographic information and clinical characteristics of the replication sample are presented in the Supplement.

**Table 1.**
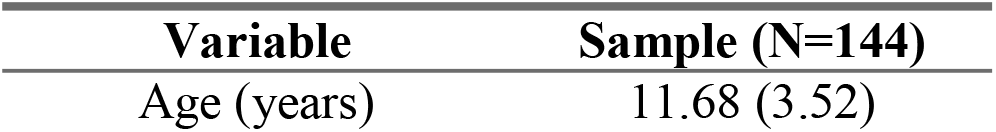
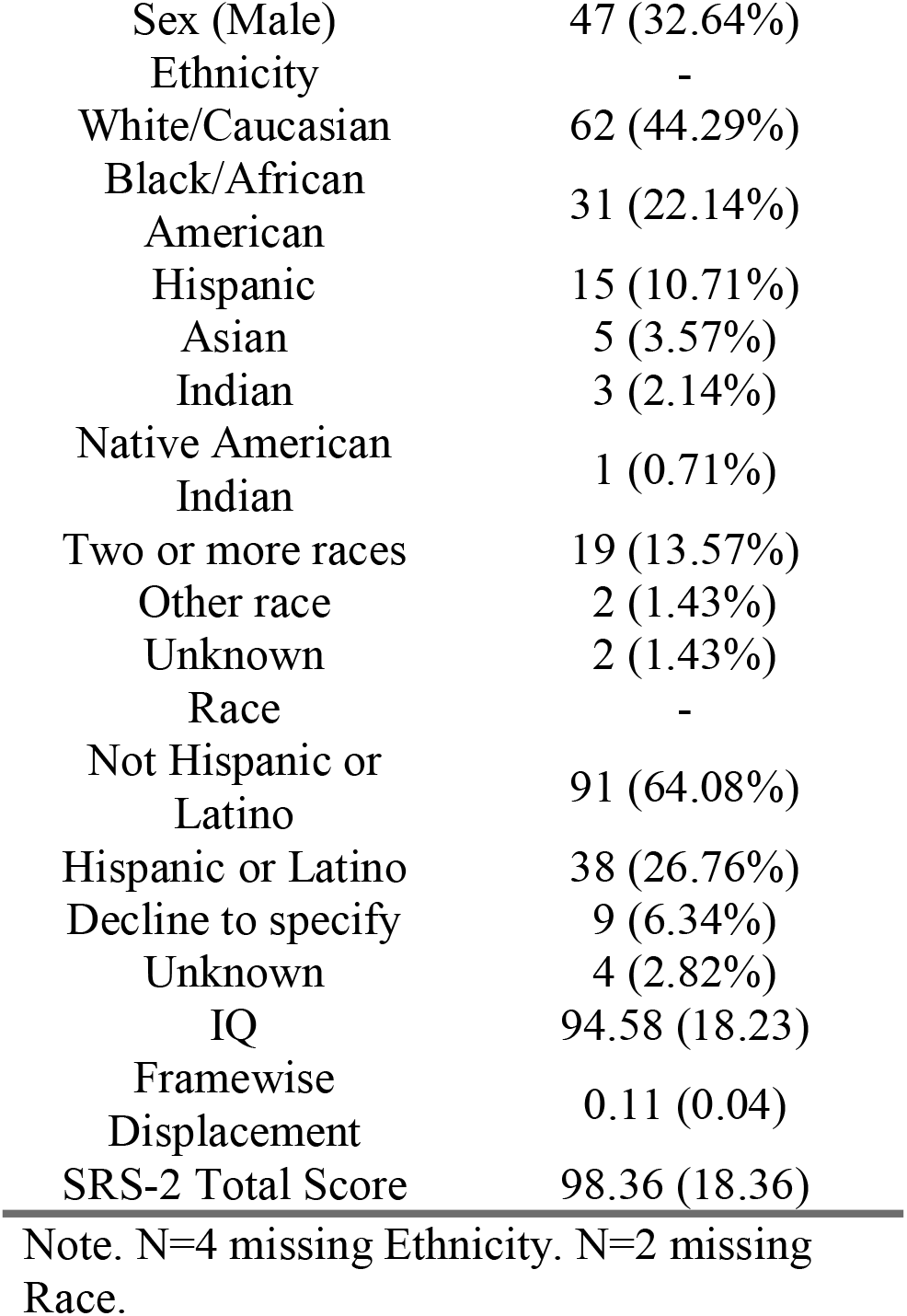
Demographic information for the Healthy Brain Network sample.

**Table 2.**
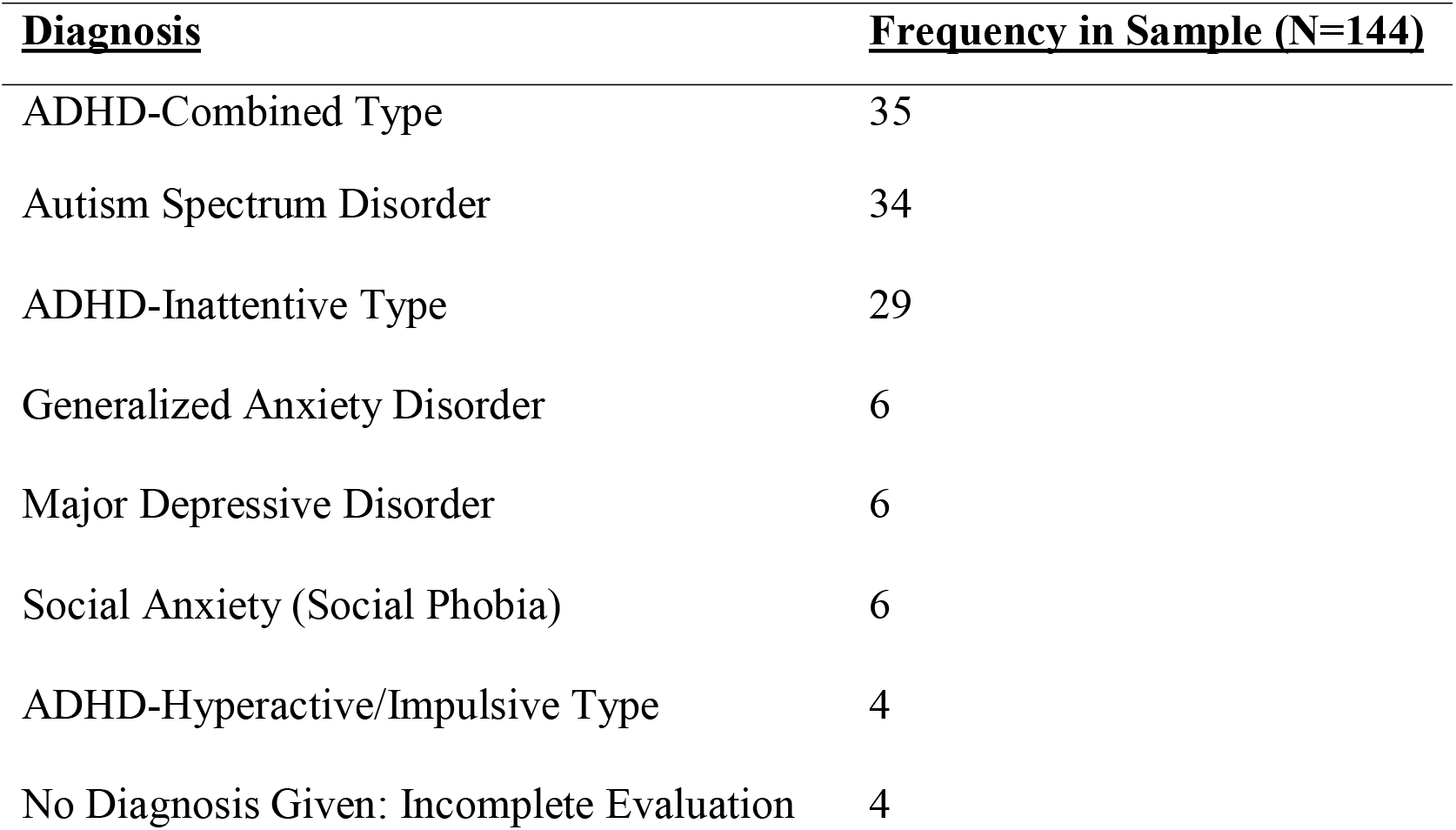
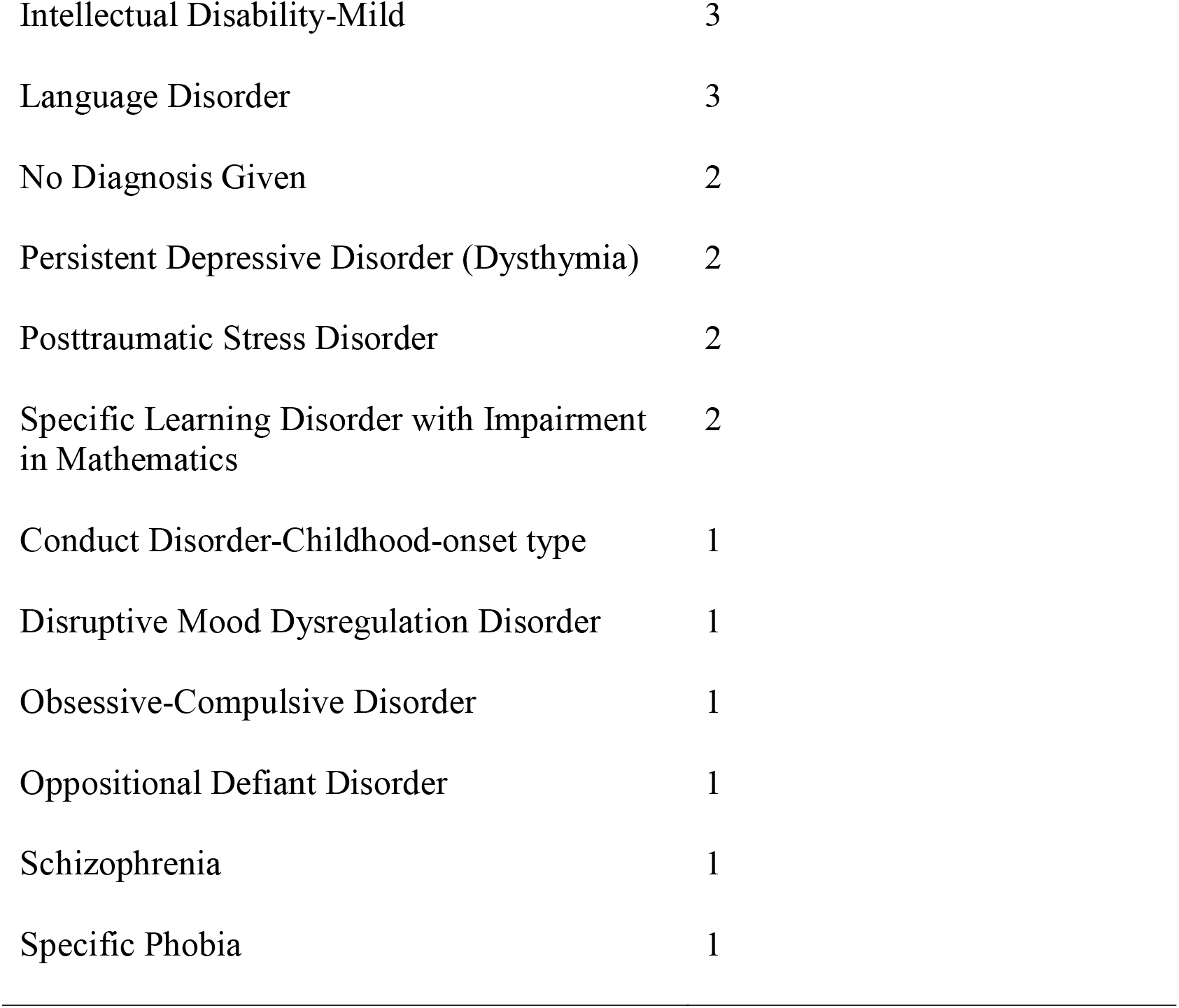
Clinical characteristics of the Healthy Brain Network sample.

### Transdiagnostic Prediction of SRS-2

As hypothesized, functional connectomes were able to predict SRS-2 total scores in the HBN sample (median *q*^*2*^=0.32, *r=*0.58, *ρ*=0.69, *p*<.001, permutation testing, 1000 iterations) (see **Figure 1**). Follow-up comparisons indicated similar prediction performance when controlling for participant age at the time of the scan (median *q*^*2*^=0.329, *r=*0.57, *ρ*=0.69, *p*<.001, permutation testing, 1000 iterations), participant sex (median *q*^*2*^=.35, *r=*0.59, *ρ*=0.7, *p*<.001, permutation testing, 1000 iterations,), head motion (median *q*^*2*^=.33, *r=*0.58, *ρ*=0.69, *p*<.001, permutation testing, 1000 iterations), IQ (median *q*^*2*^=.33, *r=*0.57, *ρ*=0.7, *p*<.001, permutation testing, 1000 iterations), and site (median *q*^*2*^=.32, *r=*0.57, *ρ*=0.69, *p*<.001, permutation testing, 1000 iterations). For prediction of the SRS-2 subscales, all the CPM models had a *q*^*2*^*s*>.04 (*r*s>.31, *ρ*s>.28) and *ps*<.001 except for the Motivation subscale (median *q*^*2*^=-.06, *p*<.001, *r=*0.21, *ρ*=0.28, permutation testing, 1000 iterations).

**Figure 1.**
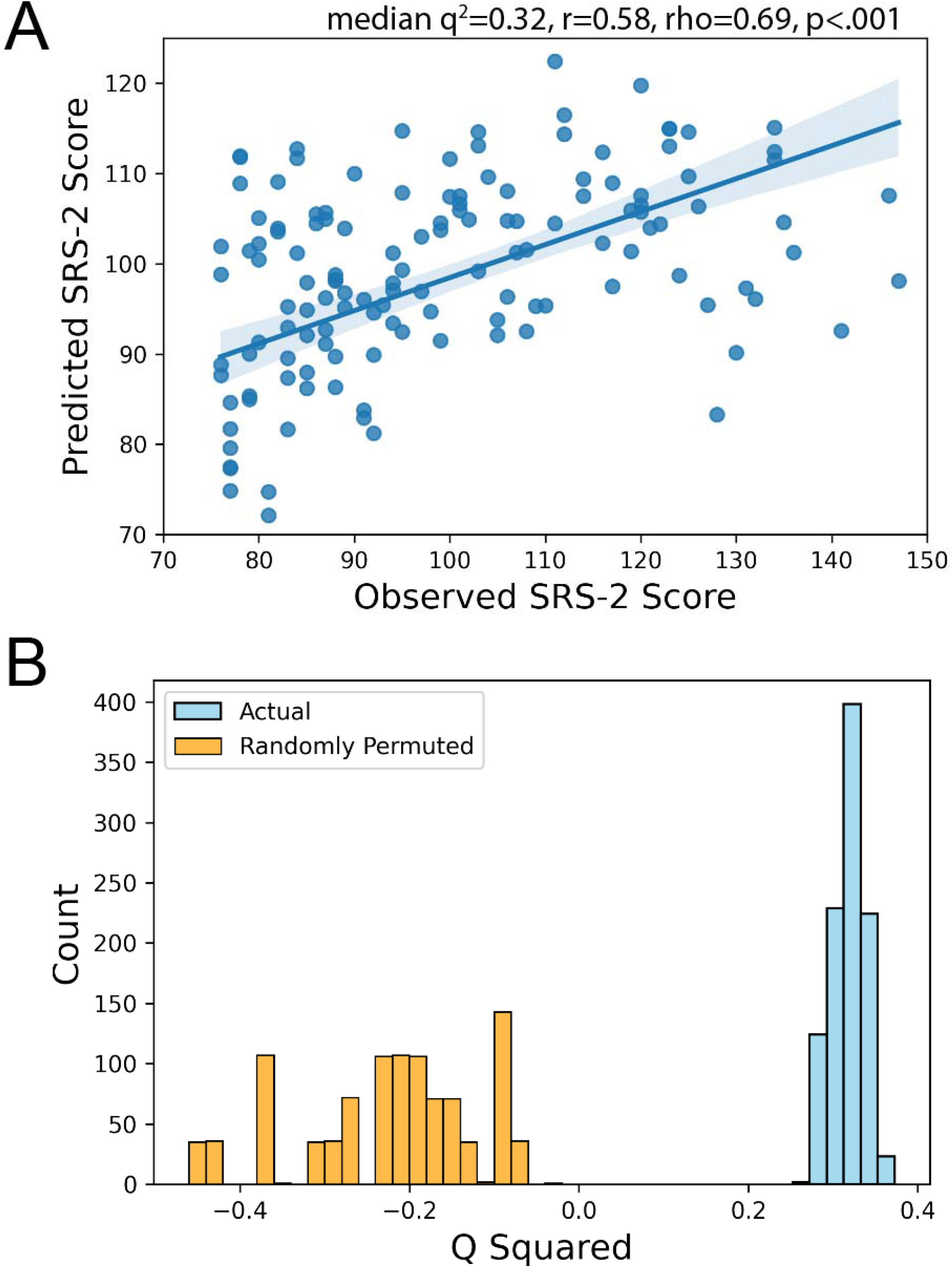
(A) Scatter plot of the association between predicted and observed scores for the connectome-based predictive model of SRS-2 scores. (B) Actual versus randomly permuted values to test the significance of the connectome-based predictive model of SRS-2 scores.

The leave-one-group-out procedure resulted in significant prediction of SRS-2 scores when individuals with an ASD diagnosis (n=59, median *q*^*2*^=.06, *p*<.001, *r=*0.38, *ρ*=0.33), individuals with an ADHD diagnosis (n=114, median *q*^*2*^=.22, *p*<.001, *r=*0.5, *ρ*=0.51), individuals with a mood disorder diagnosis (n=23, median *q*^*2*^=.229, *p*<.001, *r=*0.48, *ρ*=0.4), individuals with anxiety disorder diagnosis (n=63, median *q*^*2*^=.17, *p*<.001, *r=*0.43, *ρ*=0.43), and individuals with an “other” diagnosis (n=104, median *q*^*2*^=.35, *p*<.001, *r=*0.6, *ρ*=0.65) were left out.

Finally, there was a significant difference in network strength for the identified social impairment network between those with and without social impairment (*t*(841) = 6.65, *p* < .0001).

### Anatomical Localization

As with previous CPM studies, the resulting positive and negative predictive networks were complex and spanned multiple distributed brain regions (**Figure 2a**). While widely distributed, the predictive edges only contained a small portion of the total edges of the connectome (2166 edges in total out of 35,778, or 0.60%). Positive predictive edges of total SRS-2 scores consisted of 354 ipsilateral connections in the left hemisphere, 281 ipsilateral connections in the right hemisphere, and 418 connections between the left and right hemispheres. The negative predictive edges of SRS-2 total scores comprised 214 ipsilateral connections in the left hemisphere, 231 ipsilateral connections in the right hemisphere, and 667 connections between the left and right hemispheres. High degree nodes (**Figure 2b**) were observed for the positive predictive edges in the bilateral temporal pole, left primary sensory cortex, right orbitofrontal cortex, left hippocampus, right fusiform gyrus, and right anterior prefrontal cortex. High degree nodes for the negative predictive edges included the right inferior temporal gyrus, right pars orbitalis, left primary sensory cortex, left cerebellum, left hippocampus, left parahippocampal gyrus, and right fusiform gyrus.

**Figure 2.**
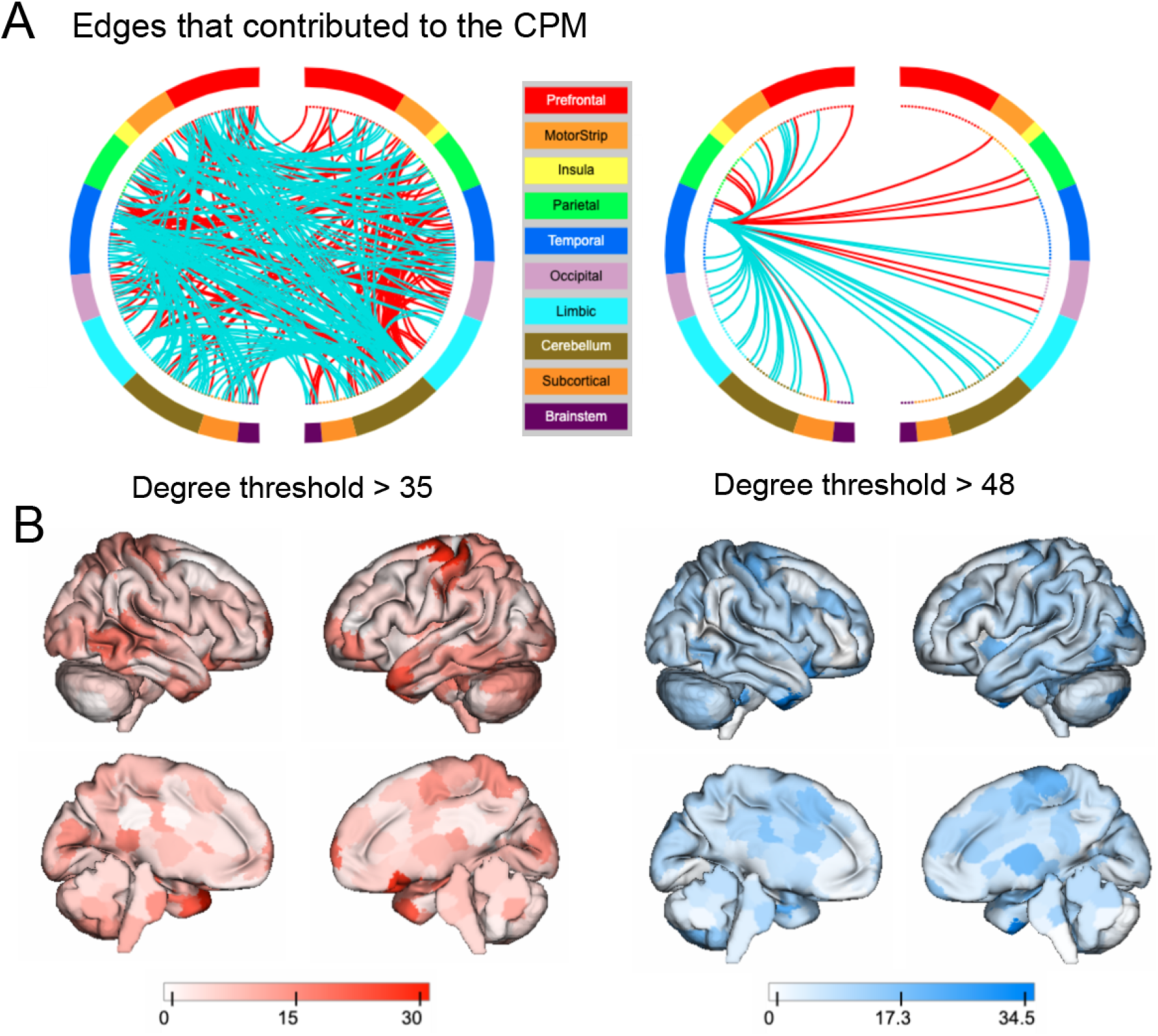
(A) Positive (red) and negative (blue) predictive networks from the CPM of SRS-2 shown at two degree thresholds for visualization purposes (threshold of 35 and 48 respectively. (B) Degree maps (unthresholded) for positive (red) and negative (blue) with darker regions indicating nodes with higher degree for the prediction of SRS-2 scores.

### Virtual Lesion Analysis

The virtual lesion analysis indicated the greatest reduction in median *q*^*2*^ was when the salience (SAL) (median *q*^*2*^=.33, *r=*0.58, *ρ*=0.69, *p*<.001, permutation testing, 1000 iterations) and subcortical (SC) (median *q*^*2*^=.33, *r=*0.58, *ρ*=0.69, *p*<.001, permutation testing, 1000 iterations) networks were lesioned (see **Figure 3**). Accordingly, the highest prediction performance for entering in single networks only into CPM was for the salience network (median *q*^*2*^=.31, *r=*0.56, *ρ*=0.69, *p*<.002, permutation testing, 1000 iterations) and subcortical network (median *q*^*2*^=.31, *r=*0.56, *ρ*=0.7, *p*<.0014, permutation testing, 1000 iterations). Next, we examined prediction performance when using only a meta-analytic social brain based upon Neurosynth (see Supplement for details). Using just the meta-analytic social brain, the prediction of the SRS-2 was significant (median *q*^*2*^=.30, *r=*0.55, *ρ*=0.58, *p*<.001, permutation testing, 1000 iterations). Finally, given our *a priori* hypothesis and the predictive power of the meta-analytic social brain, the salience network, and the subcortical network, we conducted a post-hoc test of the prediction performance if we used just the connectivity values for these networks. The performance for this model was still significant (median *q*^*2*^=.33, *r=*0.58, *ρ*=0.69, *p*<.001, permutation testing, 1000 iterations) when only using connectivity from these three networks.

**Figure 3.**
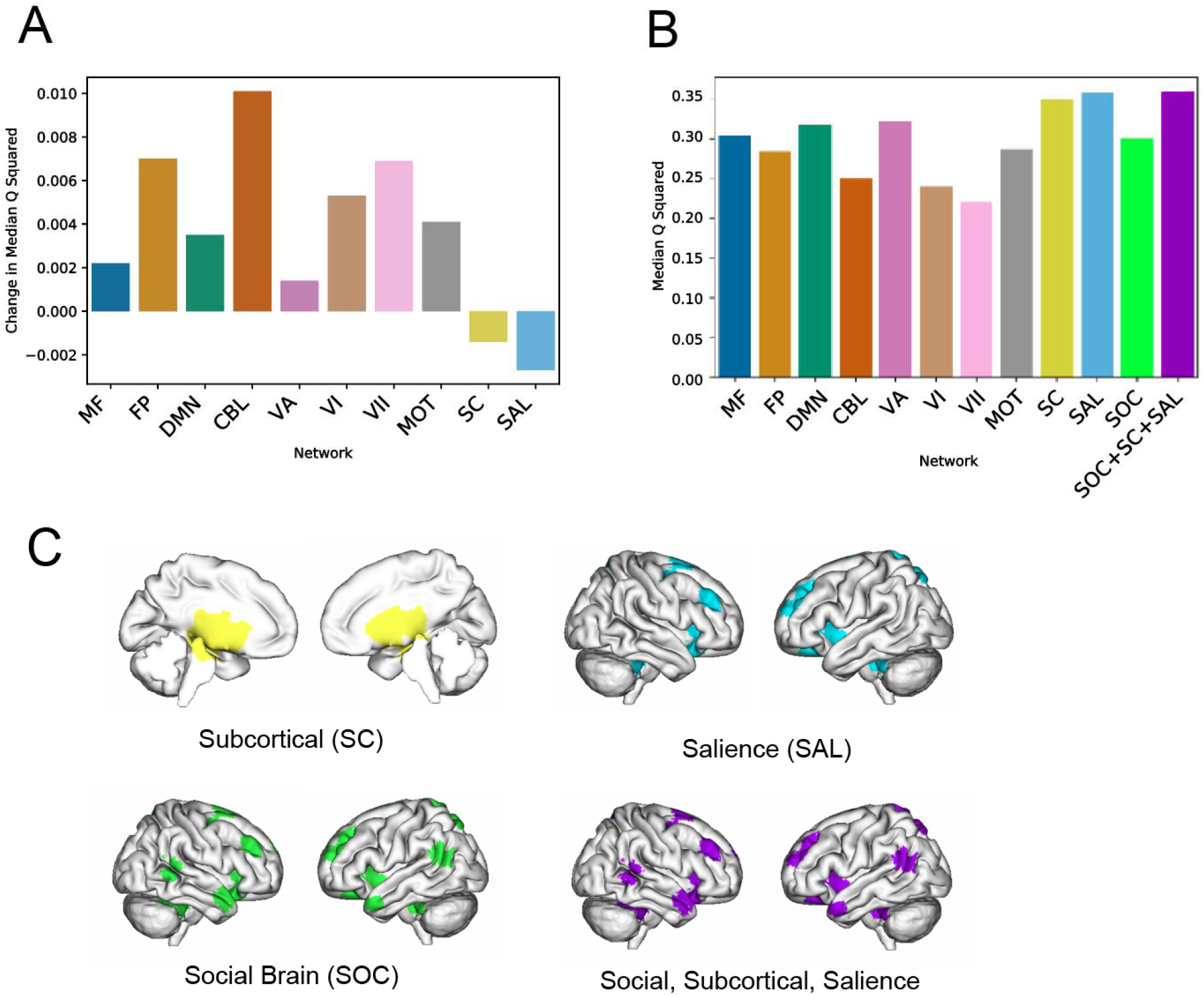
(A) Change in model performance (median *q*^2^) for when each of the canonical functional networks was virtually ‘lesioned’. (B) Model performance (median *q*^2^) when only each of the networks was used for prediction (all others set to ‘0’). (C) The 10 canonical functional networks as defined on the Shen 268 parcellation.

**Figure 4.**
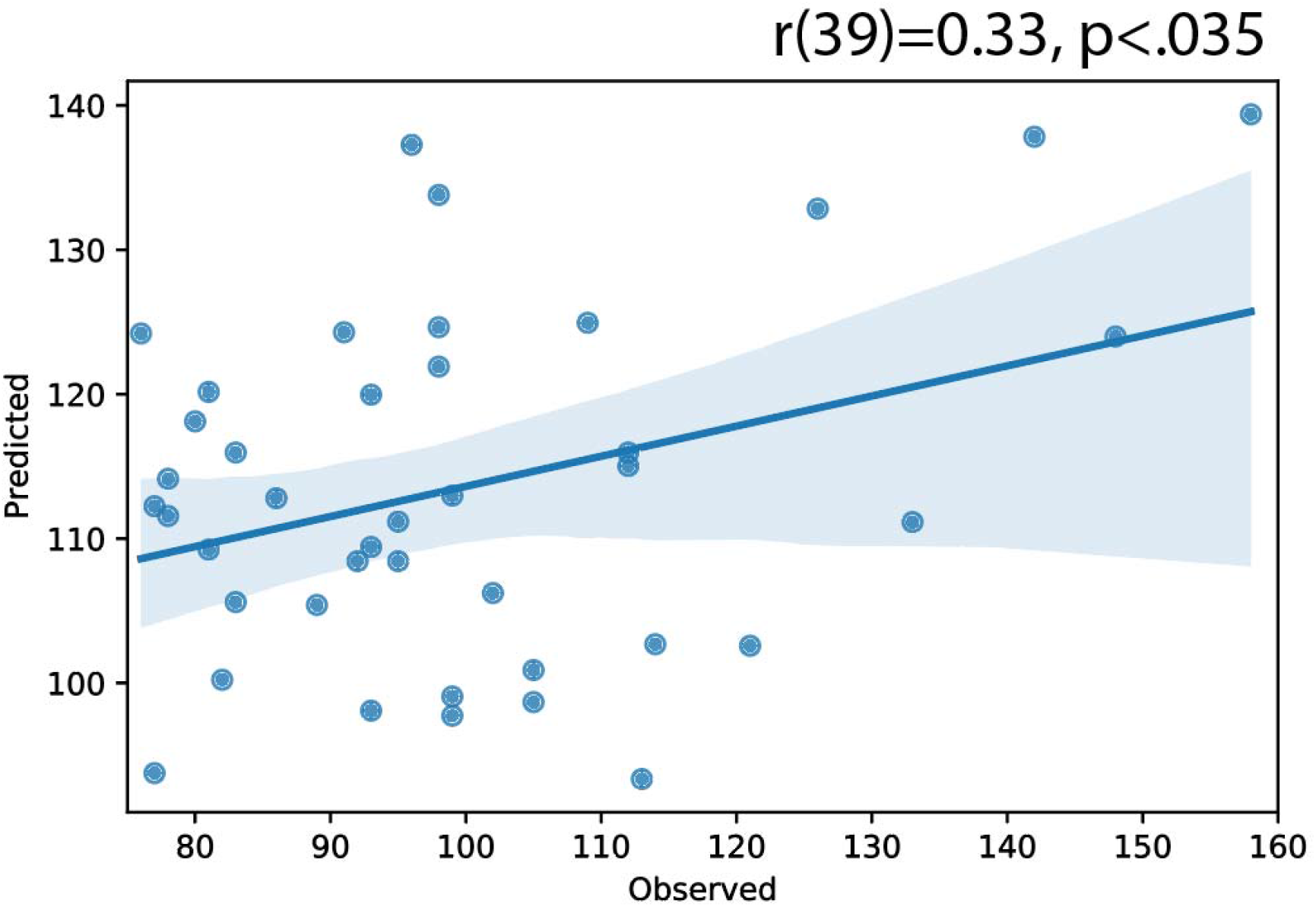
Scatter plot showing the predicted versus the observed score for external validation in the replication sample.

### Out of Sample Replication

In an independent transdiagnostic sample, the social impairment network trained in the HBN data successfully predicted SRS-2 scores *r*(39)*=*0.33, *p*<.035)

## Discussion

This study used a data-driven, whole-brain machine learning approach to identify a network of functional connectivity that predicts social impairments, transdiagnostically—a critical step as social impairments are core features of several neurodevelopmental disorders including ASD and ADHD. Critically, whereas previous investigations of the neural correlates of social impairments have focused on one or maybe two diagnostic groups, we built a model on a diverse set of participants with a multitude of psychiatric diagnoses, emphasizing the discovery of a common network of social impairment across disorders. While this common network spanned the whole brain, it was rooted in areas of the social brain, the subcortex, and the salience network, consistent with previous literature [7]. The model’s generalization across two independent neuroimaging samples is paramount for its potential utility as a transdiagnostic marker for social impairments. Altogether, we demonstrate that CPM can predict social impairments transdiagnostically across two independent samples of youths.

The current study’s findings add to literature investigating social impairments in mental health. Most prominently, our results suggest that a common network underlies social impairments in a diverse sample of youths with mental health diagnoses. While a common network was identified, alterations specific to a diagnosis likely build upon the identified network. Additionally, these results are in line with the growing number of studies that find common behavioral and neural correlations between ADHD and ASD (the two most common diagnoses across the two samples) [29-32]. When individuals with ASD diagnoses were left out of the training dataset, predictive performance was worse (*q*^2^=.06) but still significant (*p*<.001). This would be expected as social impairments are one of the defining characteristics of ASD [33, 34]. Nevertheless, both the discovery and replication samples were not limited to only diagnoses of ADHD and ASD. This diverse set of participants, as opposed to a well-controlled cohort, highlights that this model is truly transdiagnostic across different subgroups of participants.

Evidence for a CPM that can predict social impairments transdiagnostically aligns with the goals of the RDoC framework, specifically in terms of social processes [3, 35]. The SRS-2 items probe multiple aspects of social impairments including social communication, perception, and understanding of others [27, 36]. Therefore, examining functional networks that predict these impairments may be critical to understanding how atypicality in these networks may be associated with disruptions in social processes across disorders. Further, our study’s results align with the RDoC emphasis on dimensional approaches. Most brain-based predictive models for social impairments (typically for ASD) have focused on classification (does this participant have ASD or not). Predicting dimensional outcomes not only aligns with the RDoC framework but also allows prediction of subclinical symptoms and risk for developing clinical levels of symptomatology.

Our transdiagnostic model not only predicts social impairments across diagnoses but also generalizes to an independent transdiagnostic sample. The potential clinical utility of brain-based predictive models is limited unless models can generalize to independent samples. The CPM generalized to a sample that used a different functional task, a different scanner and scanning parameters, and a sample that had unique clinical characteristics. Moving forward, it will be critical to focus on the generalizability of transdiagnostic brain-based predictive models as models without external validation may be fit to sample specific noise. Models that lack external validation may be highly biased by aspects of the training data [37] and/[37],or may not generalize to samples with high degrees of co-morbidity [38]. Ultimately, clinical utility of brain-based predictive models of social impairments will be dependent on generalizability across samples, scanners, and the clinical characteristics of the sample.

Virtual lesioning highlighted the impact of the social brain, the salience network, and the subcortical network on the predictive performance of the CPM. Atypicalities in these functional networks are well-documented in disorders with high degrees of social impairments including ASD [12, 39] and ADHD [40, 41]. Further, using only connectivity values from the salience, subcortical, and social brain networks, our prediction performance was higher than when including connectivity values from across the whole brain. These finding suggest that prediction of social impairments across traditional diagnostic categories may depend on understanding the complex interactions among the salience, subcortical, and social brain networks. Virtual lesioning highlighted the impact of the social brain, the salience network, and the subcortical network on the predictive performance of the CPM. Atypicalities in these functional networks are well-documented in disorders with high degrees of social impairments including ASD [12, 39] and ADHD [40, 41]. Further, using only connectivity values from the salience, subcortical, and social brain networks, our prediction performance was higher than when including connectivity values across the whole brain. These findings suggest that the prediction of social impairments across traditional diagnostic categories will involve understanding the complex interactions among the salience, subcortical, and social brain networks. Virtual lesioning highlighted the impact of the social brain, the salience network, and the subcortical network on the predictive performance of the CPM. Atypicalities in these functional networks are well-documented in disorders with high degrees of social impairments including ASD [12, 39] and ADHD [40, 41]. Further, using only connectivity values from the salience, subcortical, and social brain networks, our prediction performance was higher than when including connectivity values across the whole brain. These finding suggest that prediction of social impairments across traditional diagnostic categories will involve understanding the complex interactions among the salience, subcortical, and social brain networks.

The Neurosynth decoding implicated associations with the model anatomy and mentalizing and the theory of mind. This finding is congruent with the multitude of studies finding that social impairments are associated with aberrant functional connectivity and activation in the theory of mind/mentalizing brain regions for ASD [42-44] and ADHD [45]. The image decoding of the negative predictive edges had the greatest overlap with the activation map of the cerebellum. The role of the cerebellum in social processes, including mentalizing, is beginning to be understood [22, 46]; further, connectivity differences have been observed in studies of individuals with social impairments such as ASD [47, 48].

The current study has several strengths, including using a whole-brain predictive modeling approach, image decoding, and virtual lesions to understand the predictive edges and test the model across two large transdiagnostic samples. However, several limitations should also be mentioned and considered. First, the replication sample size was modest (n=41); the full dataset of the replication sample was also substantially smaller than the HBN and therefore yielded a smaller number of participants with elevated SRS-2 scores. Both samples had modest sample sizes as it is difficult to obtain data from children experiencing high levels of social impairments [49, 50]. However, the cross-dataset analysis demonstrated that the discovery sample CPM could predict SRS-2 scores in a small transdiagnostic dataset of participants with elevated social impairments. Second, the study focused on SRS-2 scores of social impairments as they have been suggested as a potential transdiagnostic tool to measure impairments in the social domain [51]. Future studies will be needed to examine the edges that predicted social impairments across the independent transdiagnostic samples, generalizing to other transdiagnostic measures of social impairments in youth. Third, it is important to acknowledge that the CPM built in the study is predictive of social ‘impairment’ not of social functioning.

Therefore, the models are limited to individuals that are experiencing high levels of symptoms and may have limited utility in samples with low levels of social impairments. Lastly, the CPM was intentionally trained and tested in transdiagnostic samples; however, this may limit its generalizability to non-transdiagnostic samples. Namely, the model is not diagnosis-specific and while subset of edges from the transdiagnostic model may predict social impairments in an ASD-only sample, there may be additional ASD-specific edges that are not shared with the transdiagnostic model. Therefore, future studies will be needed to examine the common and unique edges that are predictive of social impairments transdiagnostically and those that are disorder specific.

## Conclusions

The current study demonstrates that whole-brain functional connectivity during a social movie watching task can predict social impairments within a transdiagnostic sample as well as in an independent transdiagnostic sample of youth. Networks that predicted social impairments included primarily social brain regions, including the temporal pole, inferior temporal gyrus, anterior prefrontal cortex, and regions not traditionally conceptualized as part of the social brain, including the cerebellum, hippocampus, and primary sensory cortex. These predictive edges identified in the discovery sample predicted social impairments in an independent sample. As both samples had high degrees of co-morbidity, the networks identified that predict social impairments across the sample may be critical for targeting neurobiologically informed interventions for youth experiencing social impairments that are common in several childhood neuropsychiatric diagnoses.

## Data Availability

http://fcon_1000.projects.nitrc.org/indi/cmi_healthy_brain_network/

## Declarations

### Ethics

This study was reviewed and approved by the local ethical committee (institutional review board at the Yale University School of Medicine), and it was conducted in accordance with the declaration of Helsinki.

## Acknowledgements

AJD was funded by CTSA Grant Number TL1 TR001864 from the National Center for Advancing Translational Science (NCATS), a component of the National Institutes of Health (NIH). Support was provided to DS by the NIMH grant P50MH115716. Replication sample data was obtained from the study supported by NIMH grant R01MH101514 to DGS. KI is supported by NCATS grant KL2 TR001862 and a Child Study Center Junior Faculty Development Pilot Award.

## Authorship Contributions

AJD and DS designed the study. AJD, DS, VK, JD, LT analyzed the data. AJD, DS, VK, JD, LT, KI, and DGS wrote the manuscript.

## Competing Interests

The authors declare no competing interests.

## Notes

### Competing Interest Statement

The authors have declared no competing interest.

