## Supplement for "Predicting Transdiagnostic Social Impairments in Childhood using Connectome-based Predictive Modeling"

**Supplementary Methods**

**Discovery sample.** The sample was recruited using a community-referred recruitment model. Further details about the recruitment and study design have been reported [28]. Participation in the HBN included a comprehensive diagnostic evaluation via clinician-administered assessments. Licensed clinicians conducted all diagnostic evaluations, including a semi-structured diagnostic interview: the Schedule for Affective Disorders and Schizophrenia Children’s version (KSADS) [32]. The computerized version was conducted as a semi-structured DSM-5 based psychiatric interview that includes a parent and child interview that results in automated diagnoses. After administration of all interviews, all materials collected during study participation were included in the consensus DSM-5 diagnoses by the clinical team [28].

**Replication sample.** For the full replication sample, 18 participants had an ASD diagnosis which was confirmed by either the Autism Diagnostic Interview-Revised [33] or Autism Diagnostic Observation Schedule-2^nd^ edition [34]. Recruitment for this transdiagnostic sample occurred at an outpatient child psychiatry clinic at the Yale Child Study Center and outreach to local schools and mental health providers (see Supplement Table 1).

**Supplement Table 1. Demographic and clinical characteristics of the Replication Sample.**

| **Variable** | **Sample (N=41)** |
| --- | --- |
| Age (years) | 12.31 (2.28) |
| Sex (Male) | 34 (82.93) |
| Head Motion | 0.18(0.25) |
| SRS-2 Total Score | 99.76 (19.84) |

**Social Responsiveness Scale-2.**  The SRS-2 comprises 65 items in which items are scored on a 4-point Likert-type scale ranging from not true = 1, sometimes true = 2, often true = 3, to almost always true = 4 [34]. It includes the following subscales: Social Awareness, Social Cognition, Social Communication, Social Motivation, and Restricted Interests and Repetitive Behavior [34]. The SRS-2 calculates a Social Communication Index, the sum of the responses for the Social Awareness, Social Cognition, Social Communication, Social Motivation subscales [34]. The NIMH workgroup has suggested the SRS-2 as a potential transdiagnostic measure of social impairments [35]. Further, recent empirical studies have provided evidence that the scale adequately captures the RDoC social constructs. These include the ability to perceive and interpret social signals, motivation to engage in social interactions and form social bonds, and having the skills necessary to initiate and maintain social interactions and relationships [36, 37].

**Connectivity processing.** Each participant’s functional MRI data were transformed into common space using a concatenation of two registrations: a rigid transformation of the functional data to the high-resolution T1-weighted image and a nonlinear registration of the participant’s T1-weighted image to the template space using a previously validated algorithm. Mean frame-to-frame displacement was calculated for each functional dataset, and data with a mean frame-to-frame displacement greater than 0.2 mm was removed from the analysis (n=34). Whole-brain functional connectivity matrices or ‘connectomes’ were calculated from the functional connectivity data using the Shen-268 functional parcellation. The mean time course for each node of the parcellation was calculated by averaging the time courses across all voxels in the node. These time courses of every pair of nodes were correlated, and the correlation coefficients for each pair will be Fisher z-transformed resulting in a symmetric 268x268 connectome.

**Replication Sample Connectivity Processing.** The details of the MRI data acquisition for the replication sample have been previously reported [27, 32]. Participants were engaged in a previously validated fMRI task of emotionally expressive faces during the functional MRI acquisition. Further details of the task paradigm have been reported elsewhere [32]. Functional connectivity preprocessing and connectivity matrix calculation was identical to the discovery sample resulting in 268 x 268 connectivity matrices for each participant.

**Leave-one-site-out Cross-validation:** As the study was conducted at 3 sites, we conducted a leave-one-site-out cross-validation (3 sites in the HBN: Staten Island, Rutgers University Brain Imaging Center (RUBIC), and Citigroup Biomedical Imaging Center (CBCIC) to demonstrate that the model predicted across data acquisition sites for the HBN. For the sample, 65 participants were scanned at the Staten Island site, 9 participants were scanned at the RUBIC site, and 70 participants were scanned at the CBCIC. Each site was left out while the remaining sites were used for CPM. Fourth, we used an independently acquired dataset to test the model’s generalizability via external validation.

**SRS-2 Subscale Analysis.** In addition to conducting CPM for the SRS-2 total score, we tested if the subscales of the SRS-2 could be predicted from the functional connectivity matrices. We conducted CPM with the Social Awareness, Social Cognition, Social Communication, Social Motivation, Restricted Interests and Repetitive Behavior subscales, and Social Communication Index.

**Neurosynth Decoding Analysis.** To further characterize the results from the CPM of SRS-2 scores, we used the submitted results to the ‘Neurosynth Image Decoder’ (<https://neurosynth.org/decode/>). We created degree maps from the positive and negative predictive edges from the main CPM model of SRS-2 scores (separately). We compared the decoded image against the Neurosynth term with the highest correlation for both the positive and negative degree maps. Using the peak from this cluster, we examined the associated terms with these peaks (ignoring any anatomical labels).

**Virtual Lesion Analysis.** CPM predictive networks are typically widespread and complex, so we conducted a virtual lesion analysis. For a CPM-based virtual lesion analysis, predictive networks can be set to zero to examine the degradation in predictive performance attributed to a virtual lesion of that network [27, 36]. We conducted three versions of virtual lesion analysis. First, we iteratively set each functional network to zero and examined how this impacted the model performance as measured by *q^2^*. We conducted this virtual lesion analysis for the canonical functional networks: medial frontal (MF), frontoparietal (FP), default mode (DMN), motor (MOT), visual I (VI), visual II (VII), visual association (VA), salience (SAL), subcortical (SC), and cerebellum (CBL). Second, we analyzed with only leaving the connectivity values “unlesioned” for each network and setting the rest of the connectivity matrix to zero to examine the predictive performance of each network in isolation.

Third, we used the results from the Neurosynth automated meta-analysis for the term ‘social’ to identify a ‘social brain network.’ This analysis includes any studies in the Neurosynth database whose abstracts include the term “social” at least once. This resulted in the automated meta-analysis of 1,302 studies (**Figure X**). We thresholded the association map for the term “social” at a z-score of 3.1 and clustered the map using FSL’s ‘cluster’ tool at a threshold of 3.1 [38, 39]. We examined the overlap between the peaks of these clusters and the nodes of the Shen-268 atlas. This defined our meta-analytic “social brain.” The third virtual lesion analysis involved lesioning the social brain network and examining its effect on the model’s predictive performance.

**Network Strength Analysis.** To further examine the role of the predictive edges in social impairments, we examined group differences in network strength between the sample used for the CPM analysis (high SRS scores, above >75 raw score) and individuals from the HBN that had low “subclinical” SRS scores (<75 raw score). We 3xamined the extracted edge weights from summed network of edges from the CPM and used an independent samples t-test to test for a group difference. Of the 809 participants that had SRS scores equal to or below a raw score of 75, 110 were removed due to high motion (above threshold of 0.2 mm) resulting in 699 participants in the low SRS group.

**Supplementary Results**

As the study was conducted at 3 sites in the northeast United States, we tested if the predictive model could predict across sites using leave-one-group-out cross-validation and permutation testing (1000 iterations, one-tailed) to assess significance. This procedure indicated that SRS-2 scores could be predicted using the CPM across site: for the Staten Island site (n=65, median *q^2^*=.08, *p*<.001, *r=*0.37, *ρ*=0.4), for the Rutgers University Brain Imaging Center (RUBIC) site (n=9, median *q^2^*=.39, *p*<.007, *r=*0.69, *ρ*=0.78), and for the Citigroup Biomedical Imaging Center (CBCIC) site (n=70, median *q^2^*=.42, *p*<.001, *r=*0.51, *ρ*=0.66).

**Neurosynth Analysis.** We used Neurosynth’s Image Decoder to compare the results of the CPM to their databases of published fMRI studies. Using the positive degree map, the image decoder indicated the greatest degree of similarity with the functional pattern of the temporal pole (*r*=0.15). We further examined the ‘topics’ or ‘key words’ associated with this pattern of the temporal pole (meta-analysis maps), examining the peak of the activation pattern. The highest associations were with the keywords ‘theory of mind’ (*z*-score = 8.7, posterior probability = 0.87), ‘mental states’ (*z*-score = 7.33, posterior probability = 0.87), and ‘mind’ (*z*-score = 6.74, posterior probability = 0.82). For the negative degree map, the image decoder indicated the greatest degree of similarity with the functional pattern of the cerebellum (*r*=0.1). Using the peak of this functional pattern, the greatest associations with meta-analysis maps were for the terms ‘finger’ (*z*-score = 18.07, posterior probability = 0.9), ‘tapping’ (*z*-score = 14.63, posterior probability = 0.93), and ‘finger tapping’ (*z*-score = 14.18, posterior probability = 0.93).

**Virtual Lesion Analysis.** We conducted three versions of virtual lesion analysis [27, 36] to examine the sensitivity of the edges that predicted SRS-2 scores. First, we examined the CPM performance when iteratively setting each of 10 canonical functional networks to zero and calculating the median *q^2^* using 10-fold cross-validation. As the predictive edges were widely distributed, we conducted a second version of virtual lesioning. We retained the connectivity values for one of the 10 functional networks and set the rest of the networks to zero. We repeated this analysis for each of the 10 networks, retaining the connectivity values for each and setting the other networks to zero. The first virtual lesion analysis indicated the greatest reduction in median *q^2^* was when the salience (SAL) (median *q^2^*=.33, *r=*0.58, *ρ*=0.69, *p*<.001, permutation testing, 1000 iterations, one-tailed) and subcortical (SC) (median *q^2^*=.33, *r=*0.58, *ρ*=0.69, *p*<.001, permutation testing, 1000 iterations, one-tailed). Accordingly, the highest prediction performance for entering in single networks only into the CPM was for the salience network (median *q^2^*=.33, *r=*0.58, *ρ*=0.69, *p*<.001, permutation testing, 1000 iterations, one-tailed) and subcortical network (median *q^2^*=.33, *r=*0.58, *ρ*=0.69, *p*<.001, permutation testing, 1000 iterations, one-tailed). For the third version of the virtual lesion analysis, we examined the CPM prediction performance using only the meta-analytic social brain based upon Neurosynth. Using just the meta-analytic social brain, prediction of the SRS-2 was significant (median *q^2^*=.33, *r=*0.58, *ρ*=0.69, *p*<.001, permutation testing, 1000 iterations, one-tailed). As the salience and subcortical networks impacted prediction performance, we conducted a post-hoc test of the prediction performance of the CPM if we used just the connectivity values for the meta-analytic social brain, the salience network, and the subcortical network. The performance for this model was still significant (median *q^2^*=.33, *r=*0.58, *ρ*=0.69, *p*<.001, permutation testing, 1000 iterations, one-tailed) when only using connectivity from these three networks.

**Network Strength Analysis.** To further characterize the role of the networks in social impairments identified by the CPM, we examined the group differences in network strength for the network of regions identified by the CPM. The independent samples *t*-test indicated a significant group difference of network strength within the network of edges that predicted SRS-2 (*t*(841) = 6.65, *p* < .0001).
